# Revealing Shared Tumor Microenvironment Dynamics Related to Microsatellite Instability Across Different Cancers Using Cellular Social Network Analysis

**DOI:** 10.1101/2025.09.28.25336824

**Authors:** Neda Zamanitajeddin, Mostafa Jahanifar, Mark Eastwood, Gozde Gunesli, Mark J. Arends, Nasir Rajpoot

## Abstract

Microsatellite instability (MSI) is a key biomarker for immunotherapy response and prognosis across multiple cancers, yet its identification from routine Hematoxylin and Eosin (H&E) slides remains challenging. Current deep learning predictors often operate as black-box, weakly supervised models trained on individual slides, limiting interpretability, biological insight, and generalization; particularly in low-data regimes. Importantly, systematic quantitative analysis of shared MSI-associated characteristics across different cancer types has not been performed, representing a major gap in understanding conserved tumor microenvironment (TME) patterns linked to MSI. Here, we present a multi-cancer MSI prediction model that leverages pathology foundation models for robust feature extraction and cell-level social network analysis (SNA) to uncover TME patterns associated with MSI. For the MSI prediction task, we introduce a novel transformer-based embedding aggregation method, leveraging attention-guided, multi-case batch training to improve learning efficiency, stability, and interpretability. Our method achieves high predictive performance, with mean AUROCs of 0.86±0.06 (colorectal cancer), 0.89±0.06 (stomach adenocarcinoma), and 0.73±0.06 (uterine corpus endometrial carcinoma) in internal cross-validation on TCGA dataset and AUROC of 0.99 on external PAIP dataset, outperforming state-of-the-art weakly supervised methods (particularly in AUPRC with an average of 0.65 across three cancers). Multi-cancer training further improved generalization (by 3%) via exposing the model to diverse MSI manifestations, enabling robust learning of transferable, domain-invariant histological patterns. To investigate the TME, we constructed cell graphs from high-attention regions, classifying cells as epithelial, inflammatory, mitotic, or connective, and applied SNA metrics to quantify spatial interactions. Across cancers, MSI tumors exhibited increased epithelial cell density and stronger epithelial–inflammatory connectivity, with subtle, context-dependent changes in stromal organization. These features were consistent across univariate and multivariate analyses and supported by expert pathologist review, suggesting the presence of a conserved MSI-associated microenvironmental phenotype. Our proposed prediction algorithm and SNA-driven interpretation advance MSI prediction and uncover interpretable, biologically meaningful MSI signatures shared across colorectal, gastric, and endometrial cancers.

## 1. Introduction

Microsatellite instability (MSI) is a key molecular feature of many cancers [1, 2, 3, 4], which is caused by defects in the DNA’s mismatch repair (MMR) mechanisms. MSI-high (MSI-H) tumors typically have very high mutation burdens and prominent immune infiltration, making MSI status a powerful predictive biomarker for immunotherapy response and prognosis [5, 6]. In colorectal cancer, MSI-H is found in approximately 5% of metastatic cases and 15% of early-stage tumors, and these patients exhibit durable responses to immune checkpoint inhibitors [7]. Similarly, a large fraction of endometrial carcinomas (20–30%) are MSI-H [8], and current guidelines now recommend PD-1 blockade for advanced MSI-H endometrial cancer [3]. In gastric cancer, genomic analyses have defined an MSI-H subtype that carries a more favorable prognosis and high sensitivity to immunotherapy [9]. Guidelines such as those from ESMO mandate routine MSI testing in CRC due to its implications for immunotherapy, prognosis, and chemotherapy decisions [4].

In clinical practice, MSI is most often determined by PCR-based panels and immunohistochemistry (IHC) for the four MMR proteins (MLH1, MSH2, MSH6, PMS2) [10, 7, 11]. IHC is relatively inexpensive and widely available but can miss rare mutations or partial losses, whereas PCR (or next-generation sequencing) is more sensitive but requires high-quality tissue and is costlier [7]. Neither method is perfect, and both require extra wet-lab steps and resources. Importantly, not every patient currently gets tested for MSI despite its significance, due to resource limitations.

Recently, it has been shown that deep learning models can predict MSI directly from standard H&E-stained histology images [12, 13, 14, 15, 11, 16]. For example, Kather *et al*. demonstrated that a convolutional neural network could discriminate MSI-H from MSS on digitized gastrointestinal cancer slides [12]. Subsequent large, multicentre diagnostic studies have validated and extended these findings, demonstrating robust MSI prediction from routine whole-slide images across independent cohorts and scanner/preparation variability [17, 18, 14]. Attention-augmented and self-supervised architectures have improved performance and localization of MSI-associated morphologies, such as immune-rich and stromal patterns, in colorectal cancer [19]. Deep-learning pipelines have also been adapted and validated for endometrial cancer, supporting the notion that H&E contains latent morphological signals of mismatch-repair deficiency across tumour types [11].

More recent methodological work has emphasized clinical readiness by incorporating uncertainty estimation and Bayesian formulations to provide calibrated, per-case confidence measures suitable for triage or reflex testing [20]. A contemporary systematic review and meta-analysis confirm that, while performance varies by cohort and methodology, the body of evidence supports clinically relevant accuracy for MSI detection from H&E using deep learning [16]. Finally, the field has rapidly moved toward large pathology “foundation models” pre-trained on millions of tiles or whole-slide images [21]; models such as UNI [22], Virchow [23], and Prov-GigaPath [24] have achieved state-of-the-art results on a broad suite of downstream tasks and have demonstrated strong out-of-the-box MSI prediction performance, suggesting that foundation models will form the backbone of next-generation, generalisable biomarker prediction workflows.

Despite these advances, current deep learning MSI predictors have notable limitations. Most are trained in weakly supervised frameworks, operating on one slide (or bag of tiles) at a time and lacking interpretability. As black-box models, they offer limited biological insights and may struggle with generalization in low-data regimes. These challenges hinder clinical adoption and limit utility in real-world diagnostic workflows.

To address these limitations, we present a multi-cancer, interpretable approach for MSI prediction (Figure 1A). We use the pre-trained UNI pathology foundation model [22] to extract robust, high-dimensional embeddings from image tiles. We introduce a novel transformer-based aggregation method to integrate tile-level embeddings into a global slide-level representation. This architecture uses attention mechanisms [25] to weigh the importance and generate interpretable attention maps.

**Figure 1:**
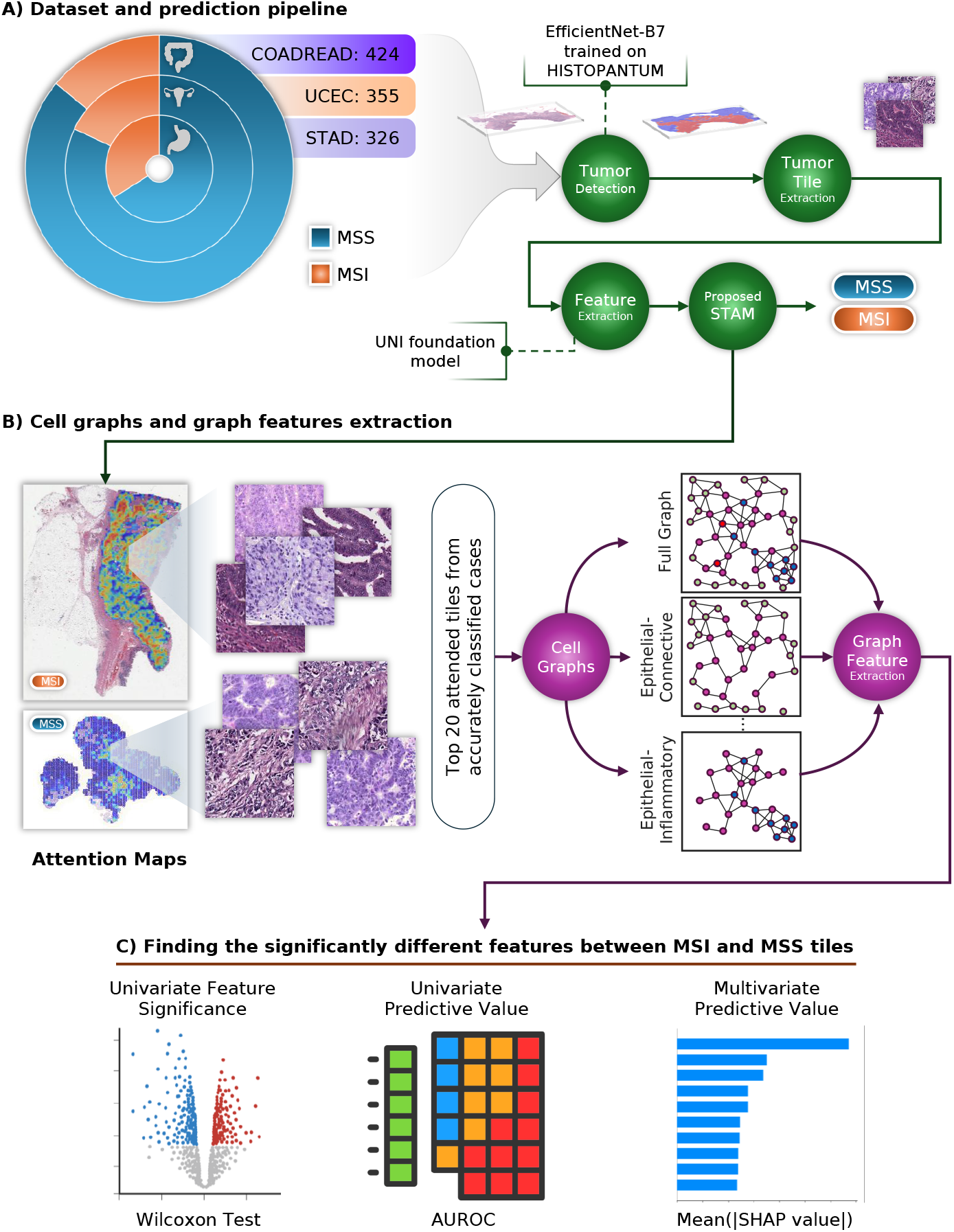
Methodology overview. (A) Dataset summary and the proposed prediction pipeline. (B) Generation of Cell Graphs in highly attended tumor tiles, and extracting graph features to describe cell-cell interactions. (C) Three categories of analyses used to find shared features related to MSIness.

Importantly, our primary objective is not only high-accuracy prediction but also understanding the tumor microenvironment (TME) dynamics associated with MSI (MSIness) across cancers. Specifically, we focus on colorectal, gastric, and endometrial cancers from TCGA to investigate whether shared manifestations of MSI in terms of cellular distribution exist across different tissue types. Although prior studies have shown immune signatures in MSI-H colorectal cancers and some studies have hinted at pan-cancer immune signatures in MSI-H tumours, detailed spatial interactions at the cellular level remain underexplored.

To interpret this shared signature, we turn to cell-graph and social network analysis of the TME (Figure 1B). We detect nuclei on H&E images and classify them (e.g. tumor epithelium, inflammatory, connective, and mitotic), then build a graph in which each cell is a node and edges connect nearby cells. In this graph representation, spatial interactions among different cell types are explicit. Such cell-graph methods have proven powerful in pathology, such as in the work of Wang *et al*., where graph neural networks applied to cell graphs can capture prognostic microenvironment patterns beyond simple counts [26]. However, in this work, we apply Social Network Analysis (SNA) metrics (e.g. degree, centrality, clustering) to these cell graphs [27, 15], providing human-interpretable measures of cellular connectivity. SNA-based features have been shown to significantly improve the prediction of molecular subtypes (including MSI) in CRC while being computationally efficient [15].

In our study, we use the attention maps from the multi-cancer predictive model to select representative tumor regions, then construct cell-graphs from those regions to quantify the TME. By comparing the SNA features and graph connectivity statistics between MSI-H and MSS cases across all three cancers using three categories of analyses (Figure 1C), we investigate whether MSI induces a conserved spatial phenotype that is common across different cancers.

The rest of this paper is organized as follows: first, the material and methods utilized in this study are explained in Section 2, then results are exhaustively investigated in Section 3 to find shared characteristics of MSIness across different cancers, and the paper is concluded with a detailed discussion on the results and the implications of this work in Section 4.

## 2. Materials and Methods

### 2.1. Dataset

The dataset used in this study was sourced from The Cancer Genome Atlas (TCGA) program [28]. Specifically, we selected a total of 1,105 cases spanning three cancer types: 424 colorectal adenocarcinoma cases (COAD-READ), 355 uterine corpus endometrial carcinoma cases (UCEC), and 326 stomach adenocarcinoma cases (STAD). MSI status for each case was obtained from [18, 12, 11], and consistent with the methodology of previous works [12, 14, 15, 18], only MSI-High cases were labeled as MSI (positive class), while both MSI-Low and microsatellite stable (MSS) cases were grouped as MSS (negative class). The class distribution between MSI and MSS cases, as shown in Figure 1A, is comparable for COADREAD and STAD, where MSI cases comprised less than 14% and 18% of the cohorts, respectively. In contrast, the prevalence of MSI was notably higher in UCEC, with approximately 34% of cases labeled as MSI; roughly double the proportion observed in the other two cancer types.

An independent external cohort from the Pathology Artificial Intelligence Platform (PAIP) Challenge was also included to evaluate the generalizability of our MSI prediction model. This dataset comprises 47 colorectal cancer cases (35 classified as MSS and 12 as MSI) sourced from three different pathology centers in South Korea.

### 2.2. Preprocessing

#### 2.2.1. Tumor detection

Since our goal is to predict MSI status from H&E-stained WSIs using only information derived from the tumor region, excluding irrelevant areas, it is crucial to first accurately localize tumor tissue. This focus allows us to specifically investigate alterations in the TME that are potentially associated with MSI, without introducing noise from surrounding non-tumor regions, such as normal tissue or background artifacts. To achieve this, we trained a binary tumor classifier using the *HISTOPANTUM* dataset [29], which comprises over 125,000 image patches labeled as either tumor or non-tumor in four cancer types, including the ones interested in this study.

For the tumor classification model, we employed *EfficientNetV2-Large* [30], a convolutional neural network architecture known for its strong performance on image classification tasks with a favorable balance between accuracy and computational efficiency. We selected this architecture due to its demonstrated ability to generalize well across heterogeneous histology datasets, particularly when trained on large-scale patch-level annotations [31].

We validated the performance of the tumor classifier on the HISTOPANTUM dataset using a leave-one-domain-out cross-validation strategy. In this setting, data from one cancer type is held out as the test set, while the model is trained on data from the remaining three cancer types. This approach mimics domain shift scenarios and assesses the model’s ability to generalize to unseen cancer types. Across all unseen folds, the model achieved a mean area under the receiver operating characteristic curve (AUROC) of 0.99±0.01 and an F1 score of 0.93±0.02, indicating strong and consistent performance. These results confirm the model’s reliability for tumor region detection in WSIs used in our study.

After training, we applied the tumor classifier to all WSIs in our MSI analysis dataset described earlier. To do this, we first performed tissue detection using the tissue segmentation model from TIAToolbox [32], which identifies tissue-containing regions on each slide and excludes background areas. Next, the trained tumor classifier was applied to the extracted tissue patches to predict the likelihood of tumor presence, resulting in a tumor map for each WSI. An example of such a tumor map is illustrated in Figure 1A.

#### 2.2.2. Patch extraction

Once tumor regions were identified, we proceeded to extract tumor image patches from each WSI to prepare the data for subsequent feature extraction and analysis. This step is essential because WSIs are extremely large, often reaching gigapixel resolution, which makes it computationally infeasible to directly process them with deep learning models in their entirety. A standard approach in CPath is to tessellate WSIs into smaller, fixed-size patches (or tiles), which can then be independently processed by neural networks for feature extraction, classification, or aggregation tasks. This patch-based strategy enables efficient computation but may cause the loss of global histological context, which is important for downstream analysis such as biomarker prediction [33]. We try to alleviate this issue with our transformer-based prediction model, which is explained later.

In our pipeline, we extracted image patches of size 224×224 pixels from tumor-containing regions at a resolution of 0.5 microns per pixel (mpp), a commonly used scale that balances spatial detail with manageable data volume. For cases with multiple WSIs available (e.g., multiple tissue sections from the same patient), we aggregated all tumor patches from the different slides into a single case-level directory to simplify downstream processing and ensure consistency across analyses.

### 2.3. MSI Prediction Model

To predict MSI status from H&E-stained WSIs, our pipeline follows a weakly supervised multiple instance learning (MIL) approach. After dividing tumor regions into small image patches, we extract high-level morphological features from them using a pretrained foundation model called UNI [22]. This model was chosen for its strong generalization across diverse tissue types and its ability to capture meaningful representations of histopathology morphology even with limited annotated data.

Patch-level features are then aggregated into a case-level representation using a novel attention-based framework called Short-Term Attention Memory (STAM). STAM captures spatial and contextual relationships between tumor regions while automatically highlighting the most informative areas that drive MSI predictions. This attention mechanism provides interpretable heatmaps, allowing visualization of tumor microenvironment patterns and potential cellular interactions associated with MSI.

A key innovation of our approach is a smart patch sampling strategy that enables simultaneous training across multiple cases, improving the model’s ability to discriminate MSI from MSS tumors while maintaining computational efficiency. By combining robust feature extraction, context-aware aggregation, and interpretable attention mechanisms, our pipeline provides both accurate MSI classification and insights into the histological patterns underlying microsatellite instability. Further technical details related to our MSI prediction can be found in Appendix A.

### 2.4. Cellular Social Network Analysis

This section outlines the methodology used to extract and analyze cell–cell interaction patterns within the tumor microenvironment (TME) using a social network analysis framework.

#### 2.4.1. Cell Graph Formation

Our approach begins by representing each tumor patch as a cell graph, where individual nuclei are modeled as nodes and their spatial relationships are represented as edges. For each extracted tumor patch, we performed nuclei detection and classification using the HoVer-Net algorithm [34], which segments nuclei and assigns them to one of five categories: neoplastic epithelial, non-neoplastic epithelial, inflammatory, connective, and dead cells. In addition, mitotic figures were identified using our in-house mitosis detection algorithm [35].

Since the tumor patches are derived from regions dominated by neoplastic epithelium, we merged neoplastic and non-neoplastic epithelial nuclei into a single epithelial category and also excluded the “dead” cell class. As a result, in each tumor patch, we focus on nuclei from four final categories: epithelial, inflammatory, connective, and mitotic.

To construct the graph, each detected nucleus is treated as a node. Edges are established based on spatial proximity to model local cell–cell interactions. Rather than connecting each node to all other nodes within a fixed radius *r*, we adopted a k-nearest neighbors (k-NN) approach: for each nucleus, edges were drawn to its 10 closest neighbors (restricted to those within radius *r*). This design choice limits the graph’s density, ensuring computational efficiency and preventing biologically implausible direct long-range connections, which are unlikely to represent direct cell–cell interactions. By constraining connectivity to the nearest neighbors, we capture the local tissue architecture while avoiding noise from distant, non-interacting cells.

##### Cell Subgraphs

To enable a more focused analysis of specific cellular interactions, we further derived cell subgraphs from the complete cell graphs (*Full-Graph*). Each subgraph isolates a subset of nodes and edges corresponding to particular biological relationships of interest. Specifically, we constructed:

- *Epithelial–Connective subgraph (EpiConGraph)*: representing interactions between epithelial and connective cells;
- *Epithelial–Inflammatory subgraph (EpiInfGraph)*: focusing on epithelial cells and their neighboring inflammatory cells;
- *Epithelial–Mitotic subgraph (EpiMitGraph)*: capturing spatial associations between epithelial and mitotic nuclei;
- *Epithelial-only subgraph (EpiGraph)*: containing only epithelial cells and their intra-class connections;
- *Inflammatory-only subgraph (InfGraph)*: retaining only inflammatory nuclei and their mutual edges;
- *Mitotic-only subgraph (MitGraph)*: including only mitotic cells and their local interactions.

Each subgraph was obtained by filtering the original full cell graph: we retained only the nodes corresponding to the relevant cell types for that subgraph and preserved the edges between them (an example is provided in Figure 1B). All other nodes and their associated edges were removed. This step allows us to disentangle the overall complexity of the TME into interpretable interaction-specific structures, enabling downstream analyses to focus on targeted biological phenomena (e.g., epithelial–immune interactions versus epithelial proliferation dynamics).

#### 2.4.2. SNA-based Features

We apply social network analysis (SNA) techniques [27, 36, 15] to quantify the structural and compositional properties of cell–cell interactions within each tumor patch. From the resulting cell graphs, we extract numerical features describing how cells are spatially arranged, how frequently they interact, and which cell types assume dominant or specialized roles within the local tumor microenvironment (TME). An overview of these features is shown in Figure 2. Since not all features are informative (as will be shown in Section 3), we do not define each one here. Instead, we outline their categories and underlying principles below, and also highlight specificly important features in the results section. A complete description of all features is provided in Appendix B.

**Figure 2:**
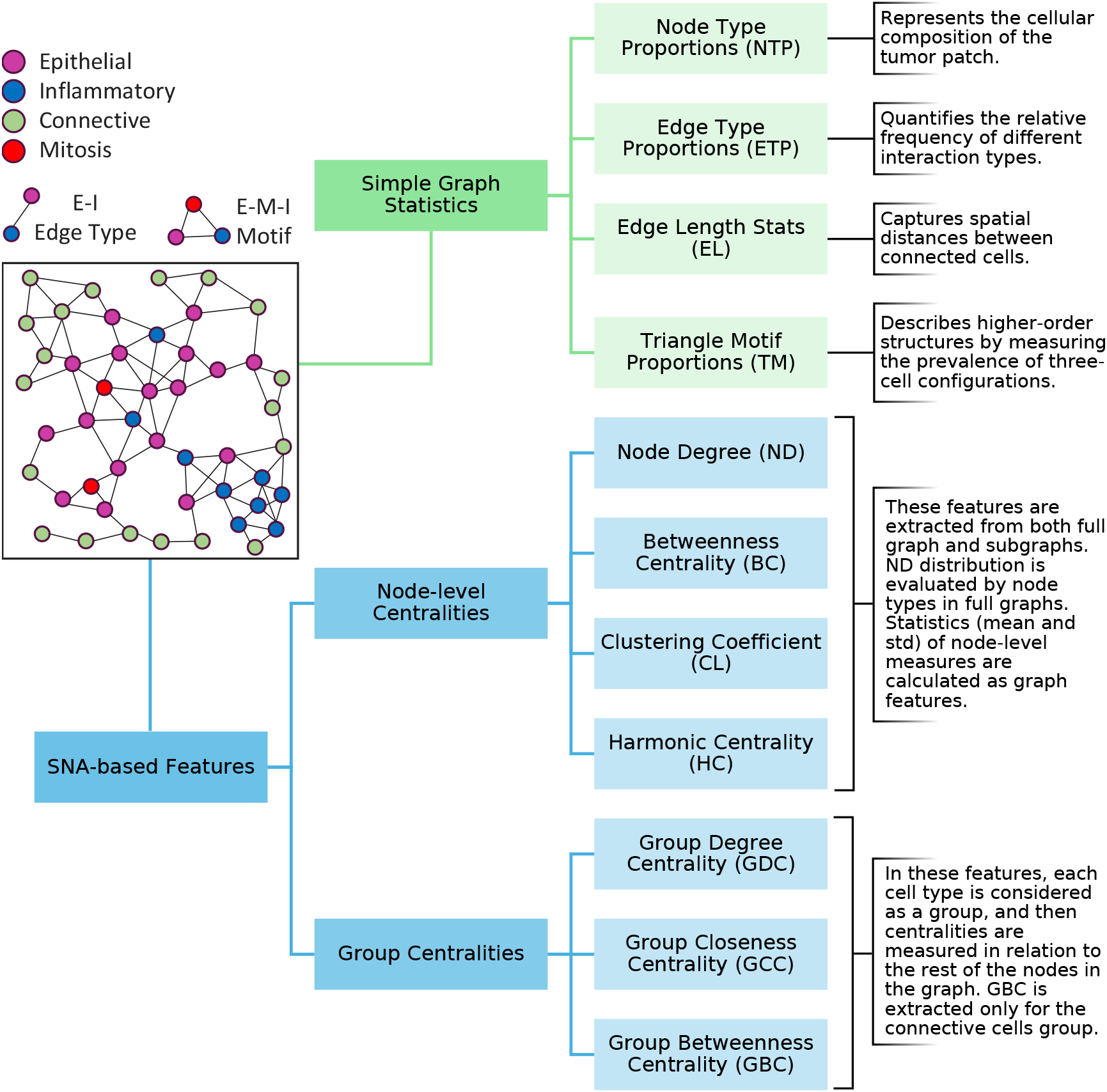
Overview of the graph features used in this study to describe cell interactions.

Our feature extraction process begins at the node level, where we compute fundamental measures such as each cell’s number of neighbors (degree) or its centrality within the network. These node-level values are then aggregated across all cells of the same type (e.g., epithelial, inflammatory), resulting in statistical descriptors such as the mean, variability, or maximum degree per cell type. At the edge level, we analyze both the types and lengths of cell–cell connections, providing insight into the frequency and spatial scale of interactions between specific cell populations.

We also calculate graph-level features, which capture higher-order organizational patterns that go beyond individual cells or single interactions. These include metrics that describe the overall cell-type composition, the prevalence of specific edge types, and the presence of small recurring structures (motifs), such as triangles, that represent clusters of three interacting cells. In addition, group centrality measures quantify the collective influence of each cell type within the entire graph, identifying which populations act as key hubs or mediators in the TME network.

Finally, we derive a set of features from cell subgraphs, which are focused portions of the full cell graph that isolate specific biological relationships (e.g., epithelial–immune or epithelial–stromal interactions). These subgraph-based metrics allow us to examine targeted cellular dynamics rather than only global properties.

### 2.5. Statistical Analyses

Our objective in these analyses is to identify cell interaction features that differ significantly between MSI and MSS cases, thereby revealing TME characteristics associated with microsatellite instability. To achieve this, we compared the distributions and predictive values of the proposed SNA-based features between MSI and MSS groups, analyzing each cancer type independently. Three complementary categories of statistical analyses were employed, each providing a distinct perspective on feature relevance.

#### 2.5.1. Univariate Statistical Testing

First, we conducted a univariate non-parametric comparison using the Wilcoxon rank-sum test (also known as Mann–Whitney–Wilcoxon: M.W.W test) [37] to determine which features exhibited statistically significant differences in their distributions between MSI and MSS cases. To ensure robust conclusions, p-values were adjusted for multiple hypothesis testing using the Benjamini–Hochberg False Discovery Rate (FDR) correction [38]. Features were considered significant if their FDR-adjusted p-values were below 0.05 (equivalently, −log(FDR) *>* 1.3) and if they showed a meaningful effect size, defined as Cliff’s delta *>* 0.15 [39]. This dual criterion ensured that selected features were not only statistically significant but also exhibited non-negligible differences in magnitude.

#### 2.5.2. Univariate Predictive Power (AUROC)

Second, to quantify and compare the direct discriminative ability of each individual feature, we calculated the AUROC using the raw feature values as univariate predictors of MSI status. Unlike the Wilcoxon analysis, which identifies statistical differences in distributions, AUROC evaluation measures the predictive strength of each feature for classifying MSI versus MSS cases. Moreover, the directionality of the AUROC (i.e., whether values are greater than 0.5 or less than 0.5) provides insight into whether higher feature values are associated with MSI or MSS cases, respectively, adding interpretability to the biological role of each feature.

To assess whether the observed AUROC values were significantly better than chance, we performed permutation testing. Specifically, MSI labels were randomly shuffled 1,000 times, and the AUROC was recalculated for each permutation, yielding an empirical null distribution. A p-value was then estimated as the proportion of permuted AUROCs that exceeded the observed AUROC in magnitude. To control for multiple hypothesis testing across features, permutation-derived p-values were adjusted using the Benjamini–Hochberg FDR procedure [38]. Features with FDR-adjusted permutation p-values below 0.05 were considered to exhibit statistically significant predictive power.

#### 2.5.3. Multivariate Analysis (XGBoost + SHAP)

Third, we performed a multivariate analysis to account for interactions between features. Specifically, we trained an XGBoost classifier [40] using the full feature set on 80% of data in each cancer to predict MSI status within each cancer type in the remaining 20%. This model achieved an average AU-ROC of approximately 0.75 across all cancer types (SD 0.001), indicating that the extracted features collectively carry substantial predictive information. To interpret the contribution of individual features within this multivariate framework, we employed SHapley Additive exPlanations (SHAP) [41], which provides a measure of each feature’s average impact on the model’s output across all cases. For each cancer type, we ranked features by their mean absolute SHAP values and selected the top ten features, thereby identifying those with the highest influence on MSI prediction when considered in combination with others.

These three complementary analyses (statistical testing for distributional differences, univariate predictive evaluation, and multivariate feature attribution) allow us to identify: (i) features that differ significantly between MSI and MSS cases within each cancer type; (ii) features whose importance is consistent across cancer types for each analysis method; and (iii) a subset of robust features that show significant differences regardless of cancer type or analysis approach, representing common hallmarks of MSI-associated TME.

## 3. Results

### 3.1. STAM Accurately Predicts MSI Status Across Cancers

We evaluated the performance of our proposed MSI prediction pipeline, based on the STAM aggregation method, using the TCGA dataset introduced in Section 2.1. A 5-fold patient-stratified cross-validation strategy was employed, ensuring that no patient’s data appeared in both training and validation sets. Each fold was repeated three times with different random seeds to ensure stability and reproducibility. Importantly, this evaluation was conducted in a pan-cancer setting, where data from all three cancer types (COADREAD, STAD, UCEC) were combined for both training and evaluation. The results are summarized in Figure 3A.

**Figure 3:**
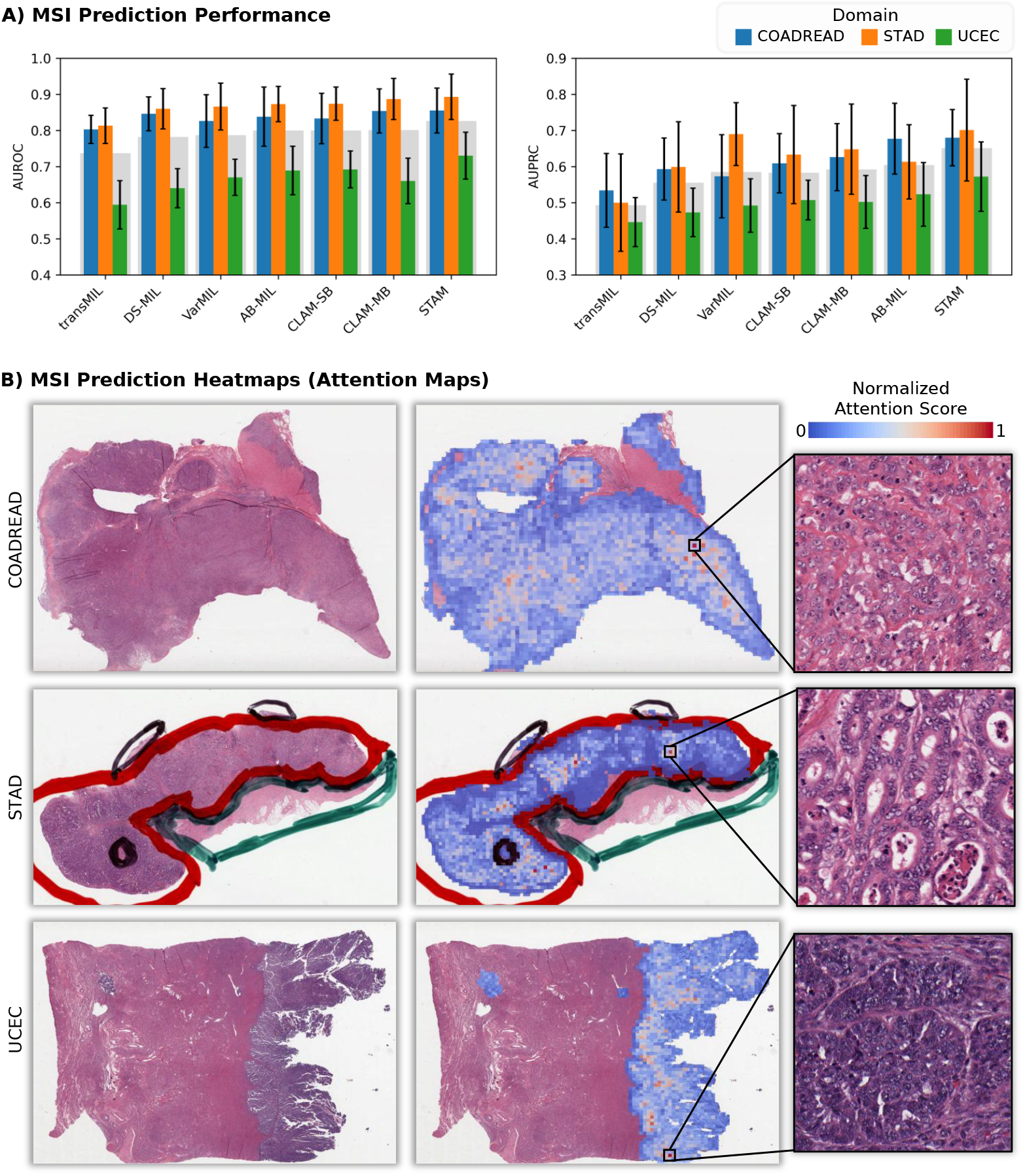
MSI prediction results. (A) Bar plots comparing the proposed method (STAM) with other recent patch aggregation methods in terms of AUROC and AUPRC. The gray bar represents the average across all three cancers. (B) Attention heatmaps that guided STAM for MSI prediction in three MSI cases. More examples are available at https://tiademos.tia.warwick.ac.uk/bokeh_app?demo=MSI.

Our STAM model was benchmarked against six state-of-the-art feature aggregation methods integrated into the same pipeline: TransMIL [42], DS-MIL [43], VarMIL [33], AB-MIL [25], CLAM-SB, and CLAM-MB [44]. These methods, briefly described in Appendix A.3, represent widely used strategies for aggregating patch-level features into case-level predictions in computational pathology.

Across all experiments, our STAM model consistently outperformed the competing methods in terms of AUROC, both per cancer type and on average. Specifically, STAM achieved AUROC scores of 0.86 ± 0.06 for COAD-READ, 0.89 ± 0.06 for STAD, and 0.73 ± 0.06 for UCEC, leading to a mean AUROC of 0.83 across all domains. This performance is higher than the next best models; AB-MIL and CLAM-MB; both of which reached an average AUROC of 0.80.

In terms of precision-recall performance, our method also demonstrated a clear advantage. The STAM model achieved an average AUPRC of 0.65, outperforming AB-MIL (0.60) and CLAM-MB (0.59). The most notable improvement was observed in the STAD cohort, where STAM achieved an AUPRC of 0.70 ± 0.10. This improvement in AUPRC highlights the model’s enhanced precision and recall trade-off in identifying the minority MSI class; a particularly important metric in imbalanced classification problems such as MSI prediction, where false positives and false negatives carry significant clinical implications.

To further test the generalizability of our approach, we evaluated the trained STAM model on the external PAIP colorectal cancer cohort, which includes MSI/MSS cases from three independent South Korean centers. Without retraining or fine-tuning, our model achieved an AUROC of 0.99 and an AUPRC of 0.98 on this cohort, demonstrating its robustness and strong transferability to datasets originating from unseen institutions.

We also investigated the impact of domain diversity in the training set by comparing the pan-cancer training strategy with a domain-specific approach, where cross-validation was performed independently within each cancer type. In the domain-specific setting, STAM’s average AUROC across the three cancer types decreased from 0.82 to 0.79 (COADREAD: 0.81 ± 0.05, STAD: 0.84 ± 0.06, UCEC: 0.71 ± 0.05). The decline was more pronounced in AUPRC, which dropped from 0.65 to 0.59 on average. These results indicate that training on data from multiple cancer types improves the model’s generalization.

### 3.2. Univariate Significance Testing (Wilcoxon Analyses)

The results of the univariate Wilcoxon rank-sum tests are presented in Figure 4. In particular, Figure 4A displays volcano plots for each cancer type, where the x-axis shows Cliff’s delta (effect size) and the y-axis shows the −log_10_(FDR) value for each feature. Features that are both statistically significant (FDR *<* 0.05) and have a meaningful effect size (Cliff’s delta *>* 0.15) are highlighted in blue. These features mark significant differences in cell–cell interaction profiles between MSI and MSS cases within each cancer type.

**Figure 4:**
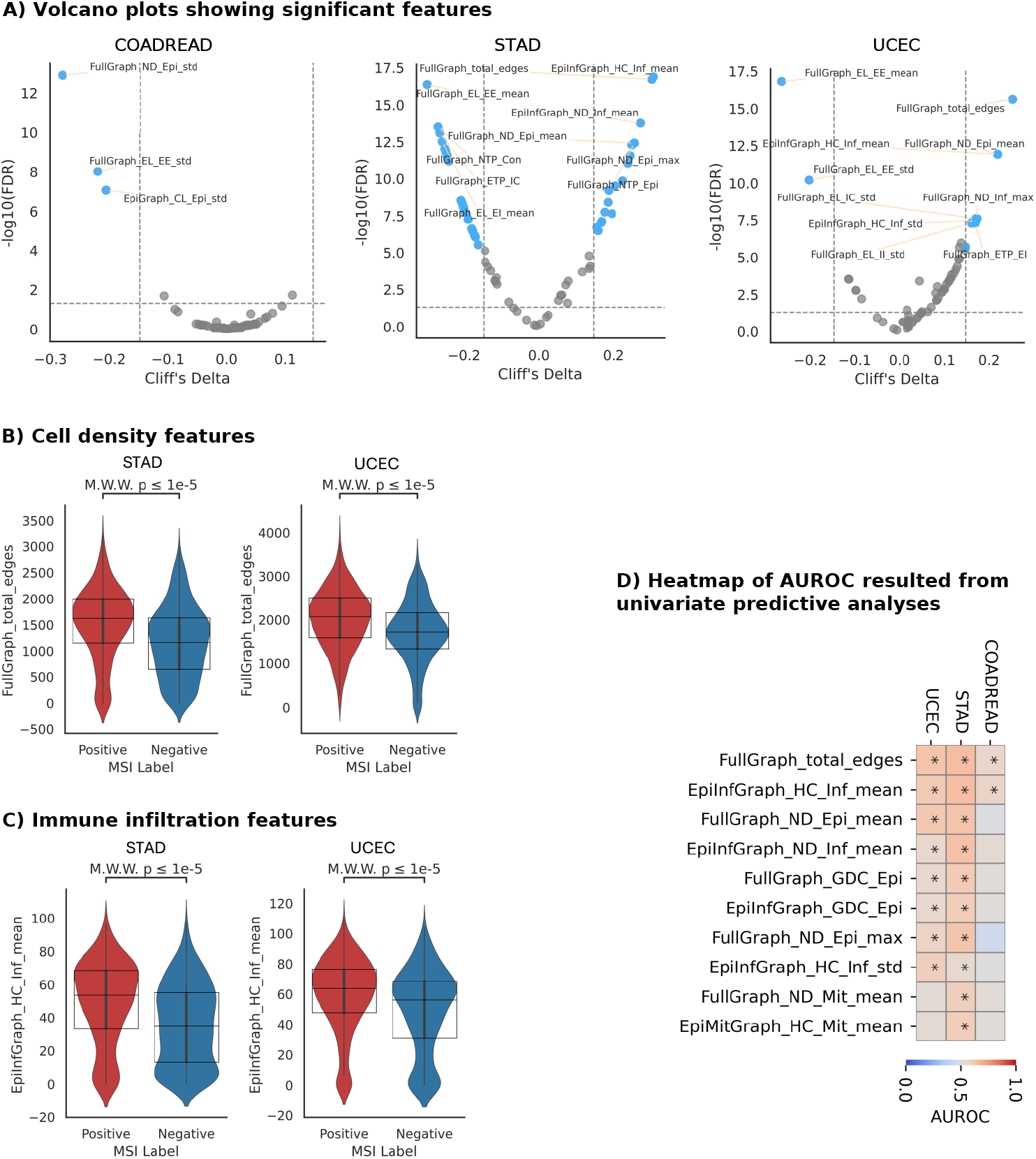
Univariate analyses results of SNA-based features for distinguishing MSI and MSS cases. (A) Volcano plots for each cancer type (COADREAD, STAD, UCEC), showing the statistical significance and effect size from the Wilcoxon test. (B) and (C) showing violin plots comparing the distribution of *FullGraph-total-edges* and *EpiInfGraph-HC-Inf-mean* between MSI and MSS cases in STAD and UCEC cohorts, respectively. The annotated *M*.*W*.*W. p* refers to the p-value from the Mann–Whitney–Wilcoxon test. (D) Heatmap of AUROCs results from univariate predictive analyses. Asterisks denote the permutation test’s corrected p-value (FDR) lower than 0.05.

#### 3.2.1. COADREAD

In this cohort, three features met our significance criteria:

- *FullGraph-ND-Epi-std:* Standard deviation of the node degree of epithelial cells. This captures heterogeneity in how epithelial cells connect to others within the tumor microenvironment.
- *FullGraph-EL-EE-std:* Standard deviation of edge lengths between epithelial–epithelial (EE) pairs. This reflects variability in spatial proximity among tumor cells.
- *EpiGraph-CL-Epi-std:* Standard deviation of clustering coefficients of the epithelial cells, which indicates how much epithelial cells are clustered.

In all features above, higher values were associated with MSS cases, suggesting that epithelial cells in MSS tumors display more heterogeneous connectivity and spatial distribution. While these features indicate variation, they do not explicitly inform whether MSI/MSS tumors are denser or more dispersed. However, insights from other cancer types help contextualize these findings.

#### 3.2.2. STAD

A broader set of features showed significant differences between MSI and MSS cases in STAD cohort. Features with *positive* Cliff’s delta values (higher in MSI cases) include:

- *EpiInfGraph-HC-Inf-mean:* Mean harmonic centrality of inflammatory cells in the epithelial–inflammatory subgraph. This suggests that inflammatory cells are more centrally located and can more efficiently communicate with epithelial cells in MSI cases.
- *EpiInfGraph-ND-Inf-mean:* Mean degree of inflammatory cells, higher values indicating more frequent interactions with epithelial cells in MSI cases.
- *FullGraph-NTP-Epi:* Proportion of epithelial cells, reflecting a higher epithelial composition in MSI tumor patches.
- *FullGraph-total-edges:* Total number of edges in the cell graph, implying a higher cell density and interaction rate in MSI tumors.
- *FullGraph-ND-Epi-mean* and *FullGraph-ND-Epi-max:* Mean and maximum node degrees of epithelial cells, both of which point to greater epithelial cell connectivity in MSI tumors.

Conversely, the following features had *negative* Cliff’s delta values (lower in MSI cases):

- *FullGraph-EL-EE-mean:* Average edge length between epithelial cells. A lower value implies that epithelial cells are spatially closer in MSI cases, supporting a denser tumor architecture.
- *FullGraph-EL-EI-mean:* Average edge length between epithelial and inflammatory cells, suggesting tighter spatial proximity between immune and tumor cells in MSI cases.
- *FullGraph-NTP-Con:* Proportion of connective cells, which is reduced in MSI cases, possibly indicating a reduced stromal component.
- *FullGraph-ETP-IC:* Proportion of edges between inflammatory and connective cells. A lower value in MSI cases may reflect a reorganization of the immune–stromal interaction landscape.

Taken together, these findings point to a TME in MSI-STAD characterized by increased epithelial cell density, enhanced immune cell connectivity, and reduced stromal presence.

#### 3.2.3. UCEC

The UCEC results largely mirror the observations in STAD, reinforcing the notion of shared MSI-related microenvironmental changes across cancer types. Features with increased values in MSI cases include:

- *FullGraph-ND-Epi-mean:* Higher average connectivity of epithelial cells.
- *FullGraph-total-edges:* Increased number of edges, indicating higher cell density or interaction rate.
- *EpiInfGraph-HC-Inf-mean:* Centrality of inflammatory cells in epithelial–inflammatory networks, again showing heightened immune–tumor interaction.
- *FullGraph-ND-Inf-max:* Maximum degree of inflammatory cells, highlighting a subset of highly connected immune cells in MSI cases.
- *FullGraph-ETP-EI:* Proportion of epithelial–inflammatory connections, pointing to increased immune infiltration or engagement.

Additionally, *FullGraph-EL-EE-mean* was again significantly lower in MSI cases, supporting the pattern of denser epithelial cell distribution.

Figure 4B–C show violin plots comparing the distribution of two consistently significant features;*FullGraph-total-edges* (Figure 4B) and *EpiInfGraph-HC-Inf-mean* (Figure 4C); between MSI and MSS cases in the STAD and UCEC cohorts. These plots illustrate both the distribution spread and median values of each feature within the two molecular subtypes. In both cancer types, *FullGraph-total-edges* is noticeably higher in MSI cases, indicating that MSI tumors tend to have greater cell density or more frequent cell–cell interactions. Similarly, *EpiInfGraph-HC-Inf-mean* is elevated in MSI cases, suggesting that inflammatory cells in these tumors play a more central role in the epithelial–inflammatory subgraph, which reflects enhanced immune engagement within the tumor microenvironment.

### 3.3. Univariate Predictive Performance (AUROC Analysis)

To assess the individual predictive power of each proposed feature for MSI status, we computed the AUROC scores in a univariate setting. The results are visualized in the heatmap shown in Figure 4D, where for clarity, only the top 10 features with the highest absolute AUROC values are presented for each cancer type.

Consistent with the findings from the significance testing, the most predictive features were those related to overall cell density; particularly of epithelial (tumor) cells; as well as the strength and proximity of interactions involving inflammatory cells. Features such as *FullGraph-total-edges, FullGraph-ND-Epi-mean*, and *FullGraph-GDC-Epi* showed strong associations with MSI status. The latter is the group-level degree centrality of epithelial cells, which measures the connectivity of epithelial cells as one group (or a super-node) with the other cell types in the network.

Similarly, features involving inflammatory cells, such as *EpiInfGraph-HC-Inf-mean, EpiInfGraph-ND-Inf-mean*, and *EpiInfGraph-GDC-Epi*, were also found to be predictive of MSI. These features measure how centrally located inflammatory cells are in the epithelial–inflammatory subgraph, how frequently they interact with epithelial cells, and how well epithelial cells connect to inflammatory cells as a group. In all these cases, higher feature values were associated with MSI, suggesting that immune–tumor interactions are both denser and more integrated in MSI cases.

It is also noteworthy that *FullGraph-total-edges* and *EpiInfGraph-HC-Inf-mean* were the only features that demonstrated consistently predictive value for MSI status in COADREAD. Although the AUROC values were modest, they reached up to 0.65 and 0.60 in STAD and UCEC, respectively.

Interestingly, features related to mitotic cells also emerged as weak but notable predictors in the STAD cohort. For instance, *FullGraph-ND-Mit-mean*, which measures the average connectivity of mitotic cells, and *EpiMitGraph-HC-Mit-mean*, which quantifies the harmonic centrality of mitotic cells relative to epithelial cells, both reached AUROC values around 0.60. These findings suggest that spatial organization and connectivity of mitotic cells may also carry information relevant to MSI status, albeit with more limited predictive strength compared to epithelial and inflammatory cell features.

### 3.4. Multivariate Feature Importance

To better understand our model’s decision-making process across different cancer types, we employed SHAP values [41], which provide both global and local interpretability of feature contributions. In each panel of Figure 5, the left bar plot shows the mean absolute SHAP values, reflecting the global importance of each feature across the dataset. Features with higher mean SHAP values are those that consistently contribute more to the model’s predictions. The right beeswarm plot in each panel visualizes the distribution of SHAP values for individual samples, illustrating the local effect of each feature. Each point represents a single case, with color indicating the original feature value (red for high, blue for low). This helps reveal how feature values influence model output in positive or negative directions, and captures complex, non-linear patterns that may differ across cancer types. The SHAP value summary plots, shown in Figure 5A–C, reveal that many of the most influential features align with those identified in our univariate analyses, particularly those related to epithelial and inflammatory cell density, connectivity, and spatial arrangement.

**Figure 5:**
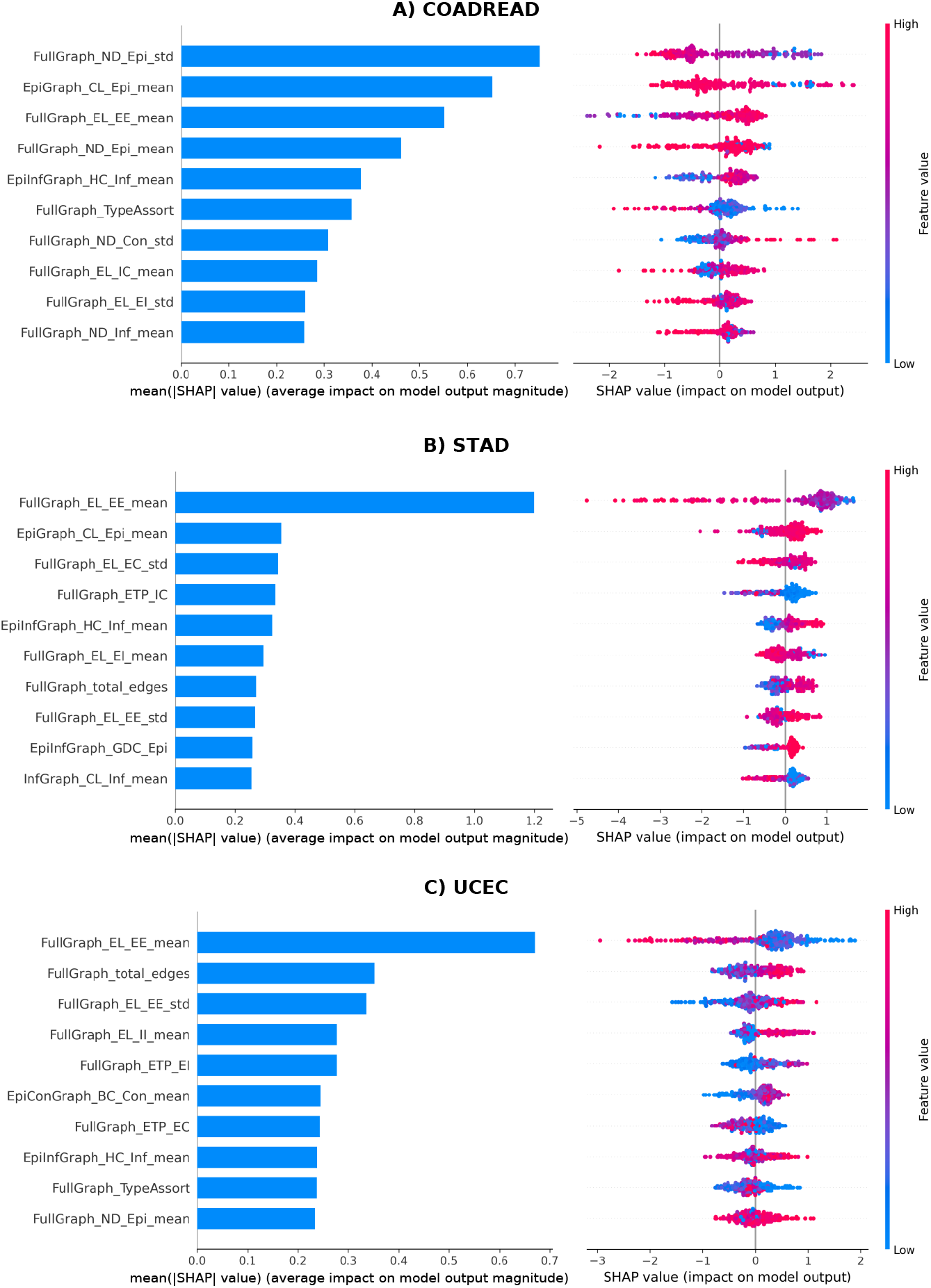
SHAP summary plots for the top predictive features in each cancer type: (A) COADREAD, (B) STAD, and (C) UCEC. Left: global feature importance. Right: beeswarm plots showing the local impact of each feature on model output.

For STAD and UCEC, features indicative of increased epithelial cell density and proximity, such as *FullGraph-EL-EE-mean* (exhibited the highest SHAP values), *EpiGraph-CL-Epi-mean*, and *FullGraph-total-edges*. These features represent the average distance between epithelial cells, their local clustering tendencies, and the overall number of cell–cell interactions in the tumor microenvironment, respectively. In both cancers, their SHAP bee swarm patterns demonstrate a clear positive association with MSI, meaning that higher values of these features increase the model’s confidence in predicting MSI. This suggests that MSI-positive tumors in STAD and UCEC tend to exhibit denser and more tightly organized epithelial cell architectures.

In contrast, for COADREAD, although the same epithelial-related features were among the top global predictors (like *FullGraph-EL-EE-mean* or *EpiGraph-CL-Epi-mean*), the directionality of their influence on MSI status was less consistent in the SHAP distributions. One feature that did stand out in COADREAD was *FullGraph-ND-Epi-std*, which measures the variability in epithelial cell connectivity. Lower values of this feature, observed in MSI cases, suggest a more uniform pattern of epithelial cell interactions; possibly reflecting a more homogeneous tumor architecture compared to MSS cases.

Features related to inflammatory infiltration also emerged as important predictors in the multivariate analysis. Notably, *EpiInfGraph-HC-Inf-mean, FullGraph-EL-EI-mean*, and *FullGraph-ETP-EI* were all associated with increased immune–tumor interactions in MSI tumors. These features reflect both the proximity and frequency of interactions between epithelial and inflammatory cells. However, features related to inflammatory-inflammatory cell interactions showed a contrasting trend. For instance, *FullGraph-EL-II-mean* and *InfGraph-CL-Inf-mean* indicated that in MSI tumors, inflammatory cells are more spatially dispersed and less clustered with one another. Biologically, this is consistent with individual immune cells in MSI tumors more actively infiltrating tumor epithelial regions and engaging with tumor epithelial cells individually, rather than forming dense immune cell clusters. This more dispersed pattern of immune cells with tumors may reflect an active anti-tumor immune response characterized by direct epithelial targeting by immune cells, rather than formation of localized immune hubs.

Interactions involving connective tissue cells also played a role in differentiating MSI and MSS phenotypes. In COADREAD, the feature *FullGraph-EL-IC-mean* showed greater edge lengths between inflammatory and connective cells in MSI cases, suggesting that immune cells are less spatially associated with connective tissue in these tumors. In contrast, STAD and UCEC tumors with MSI status exhibited lower values for *FullGraph-ETP-IC* and *FullGraph-ETP-EC*, indicating fewer connections between connective tissue cells and inflammatory or epithelial cells. Taken together, these patterns suggest that MSI tumors tend to exhibit reduced integration of connective tissue cells within tumor epithelium–immune interaction networks, possibly reflecting structural remodeling of the tumor microenvironment that favors direct tumor epithelium–immune interactions.

Another notable feature was *FullGraph-TypeAssort*, representing the assortativity coefficient of the graph based on cell type. This metric captures the tendency of cells to connect to others of the same or different types (i.e., homophily versus heterophily). This feature appeared among the top contributors in both COADREAD and UCEC, though its directional association with MSI was less consistent. In COADREAD, MSI cases tended to exhibit lower type assortativity, suggesting that cells were more likely to interact with others of the same type, which is in line with the observation made for *FullGraph-ND-Epi-std* feature. This could reflect a breakdown of heterogeneous tissue architecture in favor of more compartmentalized cellular communities in MSI tumors.

### 3.5. Cross-Analysis Integration

Across all statistical analyses and cancer types, two major categories of features consistently emerged as strong indicators of MSI status: those reflecting the density of epithelial cells and those capturing the nature of epithelial–inflammatory cell interactions. Features such as *FullGraph-total-edges, FullGraph-ND-Epi-mean, FullGraph-EL-EE-mean, EpiInfGraph-HC-Inf-mean* and *EpiInfGraph-ND-Inf-mean* were repeatedly identified as significant in univariate Wilcoxon tests, demonstrated high predictive power in AUROC analysis, and ranked among the top contributors in multivariate SHAP evaluation. These trends were most prominent in STAD and UCEC but were also observed in COADREAD to a more limited extent. For example, it is evident in image patches in Figure 6A that the MSI case cell graph is showing more clustered epithelial cells and inflammatory-epithelial connections in comparison to the MSS case, where in the top row, nodes are showing higher node degrees and harmonic centrality measures in the full cell graph and EpiInf sub-graph, respectively. A comparison of the mean values of these features across highly attended patches of MSI and MSS cases is illustrated in the radar plots of Figure 6B.

**Figure 6:**
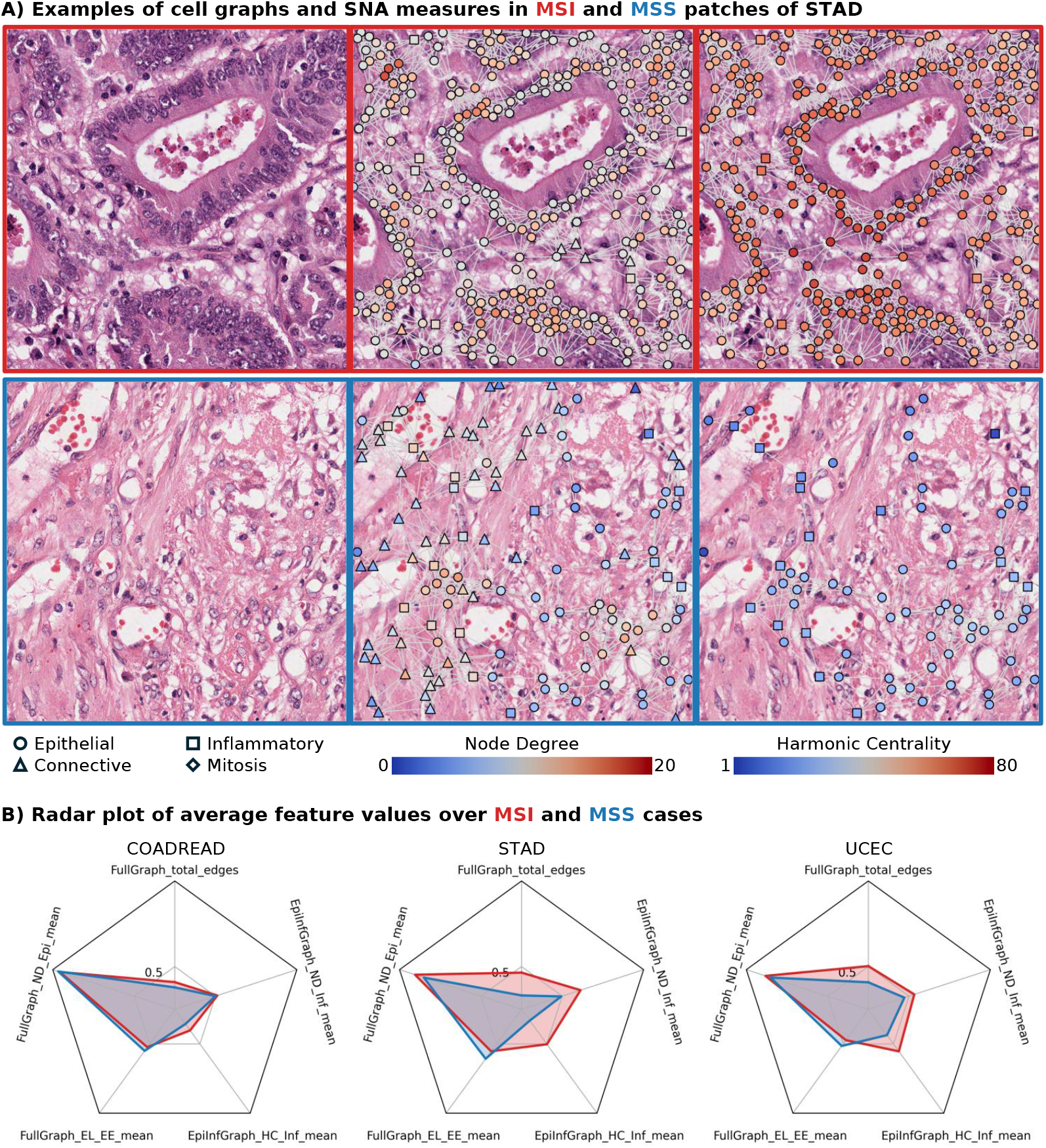
(A) Examples of highly attended patches in an MSI and an MSS case of STAD, where nodes (identified by their shapes) are colored based on Node Degrees in the full graph (middle) or Harmonic Centrality measure in the EpiInf graph (right). (B) Radar plots showing the mean values of the top 5 features identified across all analyses, calculated over the highly attended regions of all selected MSI and MSS cases.

In addition to these core categories, features describing the interaction of connective tissue cells with epithelial or inflammatory cells also appeared relevant to MSIness, particularly in Wilcoxon and SHAP analyses. For example, reduced proportions or increased spatial distances in inflammatory–connective or epithelial–connective edges were associated with MSI tumors, suggesting possible remodeling of the stromal component in the tumor microenvironment. Similar observation was reported for MSS cases of colorectal cancer, where immune cells were reported to have more interaction with connective/stromal cells [45].

Together, these results reinforce the biological relevance of tumor cellular density, immune engagement, and stromal interactions as distinguishing factors of MSI status, offering robust and interpretable markers that generalize across cancers and statistical approaches.

## 4. Discussion

Microsatellite instability is a clinically actionable biomarker in colorectal and endometrial cancers, where it plays a critical role in guiding prognosis, identifying Lynch Syndrome sufferers and guiding therapeutic decisions, including identifying patients likely to benefit from immune checkpoint inhibitors [7, 8, 46]. In gastric cancer, while MSI testing is less entrenched in standard clinical workflows, emerging evidence supports its relevance for identifying immunotherapy-responsive subgroups, particularly in advanced disease [9]. Motivated by these cross-indication implications, we hypothesized that combining MSI-labelled data from multiple cancer types could not only improve prediction performance but also reveal conserved histological features of MSIness that transcend tissue origin.

Several recent studies have established that deep learning models can accurately predict MSI status from routine H&E-stained slides, especially with the help of strong feature extractors in the field, also known as pathology foundation models [11, 15, 16, 12, 19, 21]. In our pipeline, we adopted UNI [22]; a high-capacity, self-supervised foundation model pretrained on diverse pathology data; as the feature encoder. Our focus was to use MSI prediction as a scaffold for uncovering interpretable, biologically meaningful histological patterns linked to MSIness across colorectal, gastric, and endometrial tumors.

Our proposed STAM model improves upon previous weakly supervised methods [14, 42, 25] by enabling training with multiple cases per batch—rather than one case at a time—through a smart and adaptive patch sampling strategy. By progressively prioritizing patches with high attention scores, while still maintaining diversity through random sampling, STAM enhances learning efficiency and stability, particularly in settings with limited MSI cases. The attention mechanism also produces spatial heatmaps, highlighting histological regions that most influenced the model’s predictions. These localized attention maps form the basis of our post hoc analyses, allowing us to systematically investigate and compare cellular architectures and interaction patterns between MSI and MSS cases across different cancer types.

By applying cellular graph-based analyses to the highly attended regions identified by STAM, we uncovered consistent architectural and microenvironmental trends associated with MSI. These include increased epithelial cell density, enhanced spatial and topological integration between epithelial and inflammatory cells (examples of MSI and MSS images with cell graphs overlaid are provided in Figure 6A), and distinct reorganization of the stromal compartment—findings that emerged across three different cancers and were validated through both quantitative feature analyses and expert pathologist review. As such, our study contributes not just a performant model, but a set of interpretable and biologically plausible histological signatures of MSI with potential clinical relevance.

### 4.1. Improved Predictive Performance through Efficient Training and Cross-Domain Learning

Our model consistently outperformed existing state-of-the-art weakly supervised methods across all cancer types, with particularly notable gains in AUPRC; a more informative metric than AUROC in the context of class imbalance. This improvement is largely attributable to STAM’s ability to process more diverse and representative training batches, enabled by its attention-guided patch sampling strategy. Specifically, STAM prioritizes the inclusion of highly informative patches (based on previous attention scores) while preserving variability through partial random sampling. This approach allows training with multiple cases per batch rather than one at a time, increasing both the efficiency and stability of the learning process. Importantly, the attention mechanism also produces spatial heatmaps that highlight the tumor regions driving the model’s decisions.

Importantly, we found that training STAM in a multi-cancer setting, combining slides from colorectal, gastric, and endometrial cancers, improved generalization to individual cancer domains. From a machine learning perspective, this can be interpreted as a form of implicit regularization: training on multiple domains reduces the risk of overfitting to cancer-type-specific features and encourages the model to identify more generalizable patterns. From a histopathological standpoint, MSI manifests through phenotypes such as increased lymphocytic infiltration, altered glandular architecture, and mucin production—patterns that may present with tissue-specific variations. By training on multi-domain data, the model likely gains exposure to a broader spectrum of these manifestations, allowing it to learn more transferable and domain-invariant representations of MSIness.

### 4.2. Shared Histological Features Across Cancers: Biological Insights

A major strength of our approach is its interpretability. Through a suite of univariate and multivariate statistical analyses, we identified a core set of features consistently associated with MSI status. These include metrics related to epithelial cell density (e.g., *FullGraph-total-edges, FullGraph-ND-Epi-mean*) and epithelial–inflammatory cell interactions (e.g., *EpiInfGraph-HC-Inf-mean, FullGraph-EL-EI-mean*). These patterns were particularly strong in STAD and UCEC, but also appeared to a lesser extent in COAD-READ (refer to Figure 4A and Figure 6B).

The prevalence of these features is biologically meaningful and aligns with known histological hallmarks of MSI tumors. MSI is characterized by increased tumor-infiltrating lymphocytes (TILs) [47], higher immune activation [48], and distinct epithelial architecture [1, 2]; all of which are captured in our cell graph descriptors. The stronger epithelial clustering and interaction densities we observed suggest a denser tumor core in MSI cases, while the increased epithelial–inflammatory engagement likely reflects an active anti-tumor immune microenvironment.

Furthermore, features describing reduced connective tissue integration (e.g., fewer or more distant connections between connective cells and tumor/immune cells) were also highlighted in our SHAP and Wilcoxon analyses. This may indicate that MSI tumors exhibit a form of stromal remodeling, potentially to accommodate higher immune infiltration into tumour epithelial regions, or due to modified desmoplastic response, a hypothesis that warrants further investigation.

### 4.3. Comparison to Expert Pathologist Observations

To ground our model’s findings in histopathological reality, we asked an experienced gastrointestinal and gynecological pathologist (MJA) to review attention heatmaps and top-predicted cases (without informing him of our analyses). Several of the model-identified trends were corroborated by expert review. For example, MSI tumors in all three cancer types frequently exhibited more abundant TILs and mucinous features, two canonical histological hallmarks of MSI. This aligns with the elevated inflammatory centrality and epithelial–inflammatory connectivity observed in our feature analyses.

In COADREAD, the pathologist consistently observed mucinous regions and high TIL content in MSI cases. Features such as *FullGraph-EL-EI-mean* and *EpiInfGraph-HC-Inf-mean* directly support these observations. Although traits like poor differentiation or solid/cribriform architecture were also noted by the pathologist, they were not uniformly present and did not consistently align with the learned features, reflecting their lower generalizability.

For MSS cases in COADREAD, the pathologist noted more abundant stromal regions and scarce intra-tumoral lymphocytes, which is reflected in our graph features, capturing higher connective tissue composition and fewer inflammatory interactions.

In STAD, MSI tumors again showed increased TILs and mucinous differentiation, the former captured by our graph metrics. Meanwhile, in UCEC, the most salient pathologist-identified trait was a lack of cohesion among tumor cells in MSI cases. This discohesive pattern could relate to the reduced epithelial–connective interactions observed in the SHAP results and also reported in colorectal cancer [45], although such an interpretation remains speculative.

Finally, from a multi-cancer perspective, the pathologist highlighted that while not all MSI cases displayed every hallmark feature, mucinous histology and elevated TILs were recurring motifs; a finding our quantitative analyses independently converged upon.

### 4.4. Limitations and Future Work

While our framework achieves strong performance and offers interpretable insights into MSI-related histology, several limitations warrant consideration. First, the cell detection and classification algorithms used to construct cell graphs may introduce errors or noise, especially in dense or poorly stained regions. This is particularly relevant for the COADREAD cohort, where we observed slightly weaker results, potentially due to reduced accuracy in nuclear segmentation or subtyping. Improving the reliability of these upstream steps could further enhance the quality of derived features and the robustness of downstream analyses.

Second, our feature set focused exclusively on cell–cell interactions, without incorporating direct measures of cellular morphology or tissue architecture; elements known to be histologically altered in MSI tumors. Future research could extend this framework to include morphology-informed or gland-aware features (such as the ones in [49]), offering a more comprehensive quantification of MSI-associated changes. Similarly, our analysis was conducted at the patch (tile) level, with no explicit modeling of spatial cellular heterogeneity across the whole slide. Given that MSI tumors often exhibit regionally variable immune infiltration, extending graph-based features to the WSI level could offer additional insights into how spatial context modulates MSI.

Despite these limitations, our study provides meaningful advances over the existing body of work. Unlike prior efforts that treat MSI prediction as a black-box classification problem, our framework reveals quantifiable and biologically interpretable cell-interaction features that consistently correlate with MSI status across colorectal, gastric, and endometrial cancers. To our knowledge, this is the first study to systematically uncover and validate shared histological interaction patterns of MSIness across multiple cancer types, laying the groundwork for future research into generalizable, interpretable, and clinically relevant tissue biomarkers.

## Data Availability

All data produced in the present study are available upon reasonable request to the authors.
All the data used in this work are publicly available:
TCGA: https://portal.gdc.cancer.gov/
PAIP: https://paip2020.grand-challenge.org/Dataset/
HISTOPANTUM: https://zenodo.org/records/14555794

https://paip2020.grand-challenge.org/Dataset/

https://zenodo.org/records/14555794

## Appendix A. Details of MSI Prediction Models

### Appendix A.1. Patch Feature Extraction Using the UNI Foundation Model

We selected UNI [22] for this study due to its strong generalization capabilities, high performance across diverse pathology tasks, and suitability for use in low-label settings. UNI is based on ViT-Large [50] architecture, a self-supervised model pretrained on over 100 million H&E image patches from more than 100,000 WSIs spanning 20 tissue types. It has been shown that UNI learns robust, resolution-agnostic representations, and it has consistently outperformed other feature encoders in tasks including cancer subtyping, mutation prediction, and weakly supervised slide classification, making it a strong fit for extracting meaningful features in our MSI prediction pipeline. In our pipeline, each 224×224 pixel tumor patch is passed through the frozen UNI encoder, and the resulting output is a 1024-dimensional feature vector. These features serve as compact, high-level representations of local tissue morphology and are used as input for the attention-based aggregation model described in the next section.

### Appendix A.2. Feature Aggregation and Heatmap Generation using STAM

To aggregate patch-level features into case-level representations for MSI classification, we developed a novel Transformer-based feature aggregation module called Short-Term Attention Memory (STAM). As illustrated in Figure A.7, STAM operates in two stages. First, it applies a Transformer encoder to the set of feature embeddings extracted from all tumor patches belonging to a case. This allows the model to capture contextual relationships between patches using a multi-head self-attention mechanism [51]. The Transformer outputs the same number of feature vectors—one per input patch—but each vector is now conditioned on the others, effectively embedding each patch in the context of the entire tumor region.

Next, an attention-based pooling layer is used to summarize these patch-level features into a single case-level representation. This mechanism is similar to Attention-Based Multiple Instance Learning (ABMIL) [25], where the model learns to assign higher attention weights to more informative patches. The aggregated case-level vector is then passed through a fully connected layer for binary MSI/MSS classification. The learned attention weights also provide a spatial heatmap, highlighting the regions within the slide that most influenced the model’s prediction—offering interpretability for potential clinical insights.

A key innovation of our approach lies in how we construct training batches. In standard MIL-based approaches such as ABMIL [25] or TransMIL [42], the number of patches (or instances) varies widely from case to case, and training is typically performed using one slide (i.e., one case) per batch. This is mainly due to memory constraints and the computational cost of processing all patches per slide. As a result, these models are unable to directly compare multiple cases during training, which can limit their ability to learn discriminative patterns between MSI and MSS cases.

To address this, we propose a smart patch sampling strategy (inspired by the IDaRS method introduced in [14]) that enables training with multiple cases per batch while keeping memory usage manageable. Consider the full training dataset to be **D** = {**C**_1_, **C**_2_, …, **C**_*N*_ }, where each **C**_*j*_ is the set of all patch-level feature embeddings from case *j*, and *N* is the total number of cases. At training epoch *t*, we sample a subset 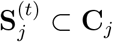, 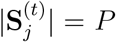 from each case in the batch. The set of case indices in the batch is **I**_*t*_ ⊂ {1, …, *N* } with |**I**_*t*_| = *B* (case-level batch-size), giving the batch:

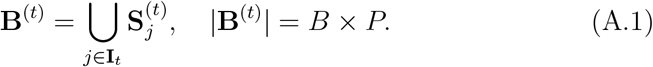

This allows the model to process *B* different cases simultaneously while keeping memory usage fixed. Initially, 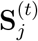 is formed via uniform random sampling from **C**_*j*_. After a warm-up phase, we bias the selection using attention scores from the previous epoch:

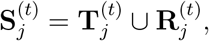

where 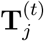 contains the top *K* = *αP* highest-attention patches from **C**_*j*_ at epoch *t* − 1, and 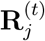 contains the remaining (1 − *α*)*P* patches sampled uniformly at random. After each epoch, the attention memory is updated for all patches in **D**, enabling the sampling process to progressively focus on the most informative regions while retaining sufficient diversity to mitigate overfitting by exploring other parts of the tumor area. Crucially, STAM enables training with multiple cases per batch, which strengthens the model’s ability to distinguish MSI and MSS by jointly learning from a broader set of inter-case variations.

### Appendix A.3. Other Embedding Aggregation Methods

Aggregation methods in computational pathology aim to combine tile-level embeddings from WSIs into a slide-level representation for downstream prediction tasks. Attention-Based MIL (AB-MIL) [25] employs fully trainable attention layers to weight the contribution of each tile embedding, enabling the model to focus on the most informative regions. CLAM-SB and CLAM-MB [44] extend the attention mechanism by incorporating a clustering-based approach: CLAM-SB is tailored for binary classification, while CLAM-MB addresses multiclass settings using attention-guided pseudo-label generation to better capture heterogeneous tissue patterns. VarMIL [33] builds on AB-MIL by adding a variance pooling module, explicitly modeling intratumoral heterogeneity to enhance performance in variable tissue contexts. DS-MIL [43] introduces a dual-stream design, coupling attention scoring with max-pooling to balance global relevance estimation and detection of the most critical instances, though it does not explicitly account for spatial correlations. In contrast, TransMIL [42] leverages a transformer-based architecture with self-attention to capture inter-tile relationships and employs pyramid position encoding to incorporate spatial context, albeit without absolute positional consistency across slides. Together, these methods illustrate the progression from simple attention-based weighting to more complex architectures that integrate heterogeneity modeling, multi-class clustering, and spatial reasoning for improved WSI-level inference. Table A.1 provides a summary of these methods and compares their main working principles.

**Figure A.7:**
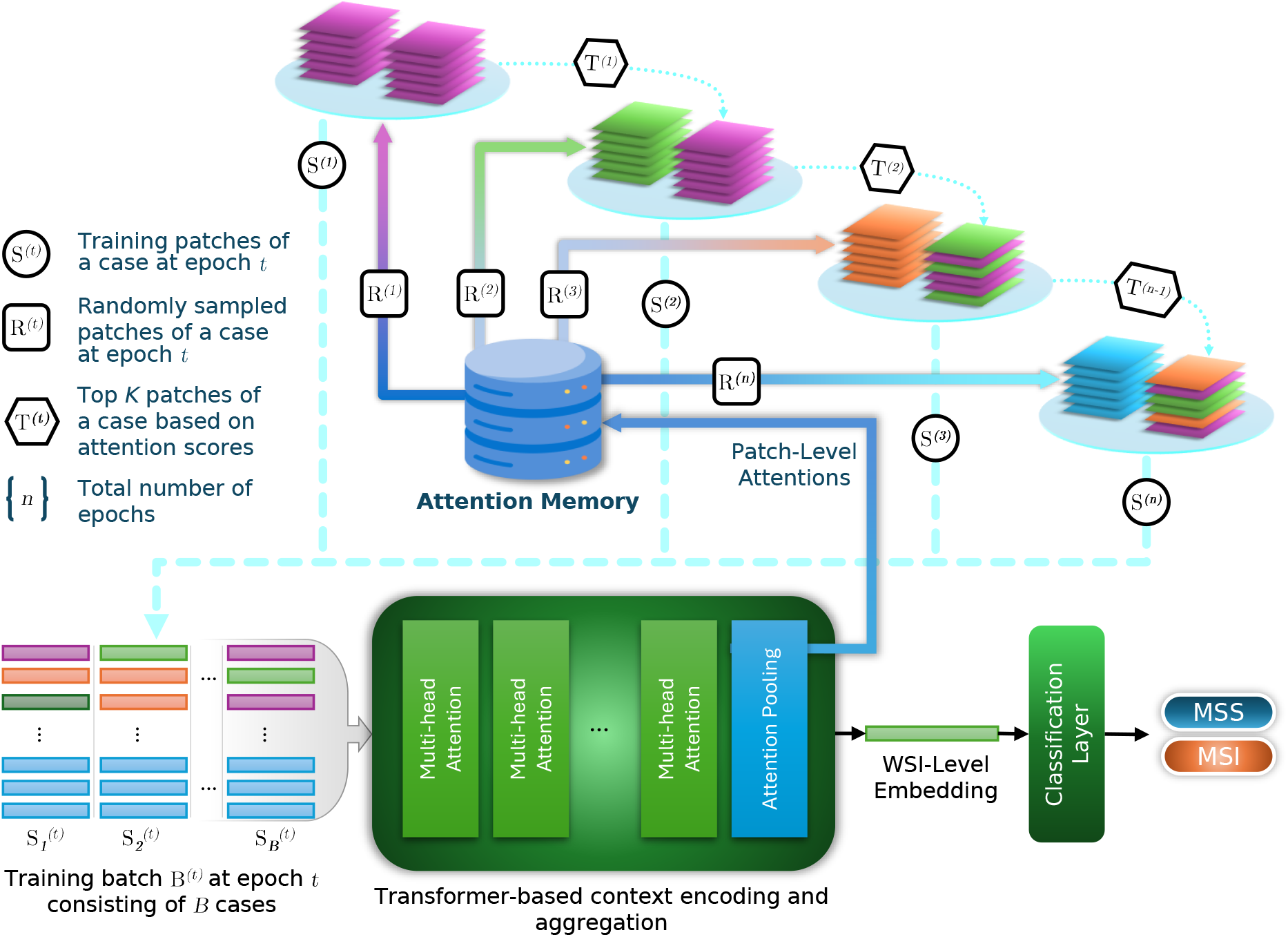
Short-Term Attention Memory (STAM) architecture for attention-based patch sampling during training and using transformer-based context encoding and WSI-level embedding aggregation.

**Table A.1:**
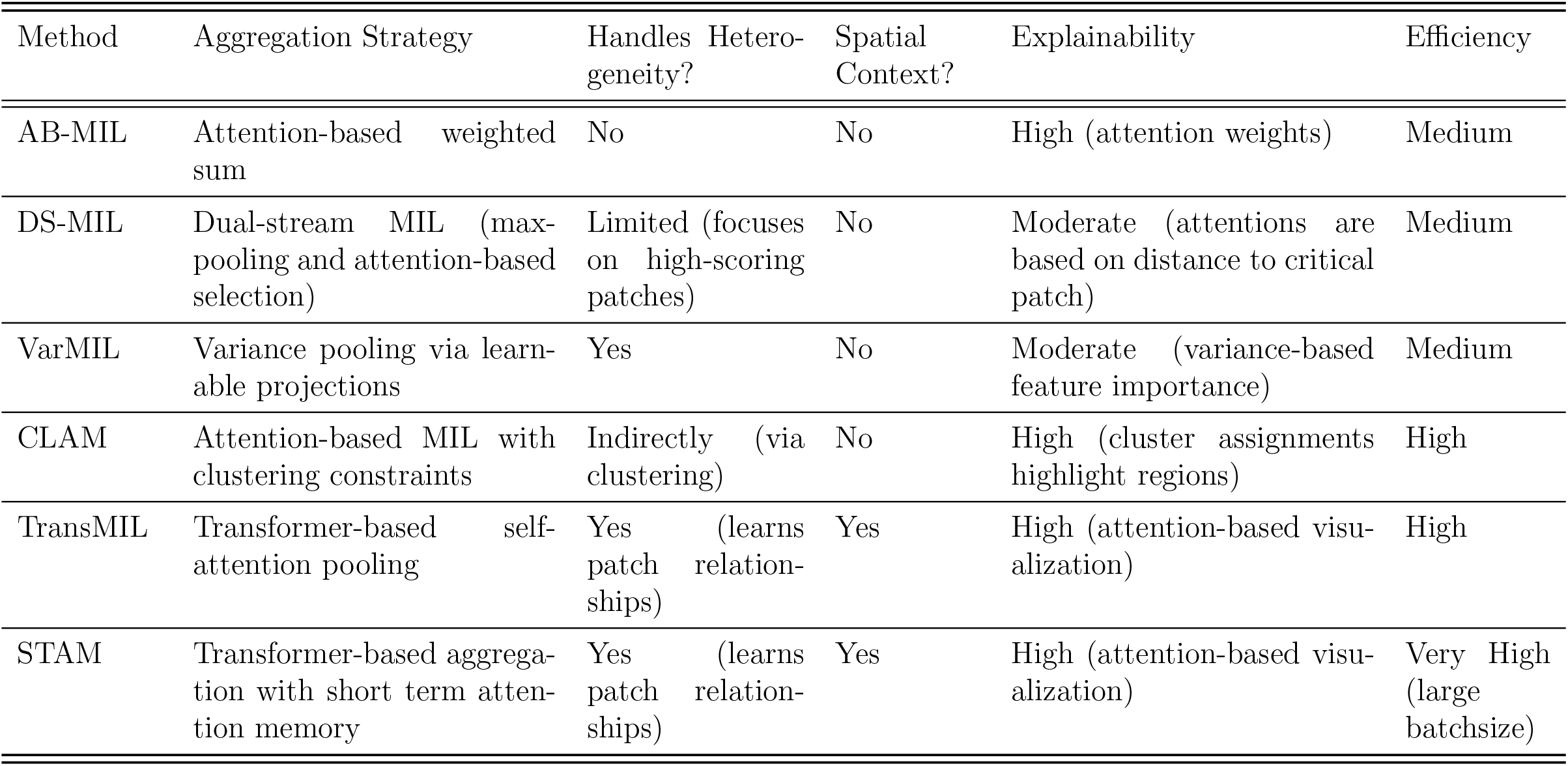
Comparing different embedding aggregation methods in CPath.

## Appendix B. Details of extracted features and their interpretation

### Appendix B.1. Degree Distribution by Node Type (ND)

Measures the connectivity of individual cells by counting the number of neighboring cells (edges) each nucleus has.

- Features: For each cell type (epithelial, inflammatory, connective, mitotic), we calculate the mean, standard deviation, and maximum degree.
- Biological interpretation: *Mean* degree reflects the average interaction level per cell type (e.g., high epithelial degree may suggest a dense tumor cell arrangement). *Variability (standard deviation)* indicates heterogeneity in interaction patterns within the same cell type. *Maximum degree* highlights the presence of hub cells that may dominate local communication.

### Appendix B.2. Node Type Proportions (NTP)

Represents the cellular composition of the tumor patch.

- Features: Proportion of each cell type and total number of nodes.
- Biological interpretation: Provides a global view of cell population balance, which may relate to immune infiltration or stromal content.

### Appendix B.3. Edge Length Statistics by Edge Type (EL)

Captures spatial distances between connected cells.

- Features: For each edge type (e.g., epithelial–epithelial, epithelial–inflammatory), compute the mean and standard deviation of edge lengths.
- Biological interpretation: Shorter mean distances may indicate tighter cellular organisation, while larger variability may reflect structural heterogeneity in the tissue.

### Appendix B.4. Edge Type Proportions (ETP)

Quantifies the relative frequency of different interaction types.

- Features: For each edge type, calculate its proportion relative to the total number of edges, along with the total edge count.
- Biological interpretation: Highlights which cell–cell interactions dominate the local microenvironment.

### Appendix B.5. Triangle Motif Proportions (TM)

Describes higher-order structures by measuring the prevalence of three-cell configurations (triangles).

- Features: For each motif type (e.g., epithelial–epithelial–inflammatory), compute its proportion relative to the total number of triangles and record the total triangle count.
- Biological interpretation: Reveals patterns of cooperative cellular arrangements that may correspond to specific microenvironmental structures.

### Appendix B.6. Group Centrality Features

Treats each cell type as a “group” node and calculates its collective centrality within the network.

- Features: Degree, closeness, and betweenness centrality for each cell type.
- Biological interpretation: Identifies which cell populations act as major connectors, are most accessible, or mediate communication across the tissue network.

### Appendix B.7. Features from Subgraphs

Derived from specialized subgraphs (Section 2.4.1) to isolate specific biological interactions.

- Features include:
  - Epithelial–Inflammatory subgraphs: mean degree, variability, group degree centrality, harmonic centrality.
  - Epithelial–Mitotic subgraphs: similar measures focusing on tumor proliferation.
  - Epithelial–Connective subgraphs: mean degree and betweenness centrality to assess stromal interactions.
  - Single-type subgraphs: clustering coefficients (mean and standard deviation) for epithelial-only, inflammatory-only, and mitotic-only networks.
- Biological interpretation: Enables fine-grained analysis of targeted interactions, such as tumor–immune engagement, stromal architecture, and local proliferative activity.

## References

[1] R. J. Hause, C. C. Pritchard, J. Shendure, S. J. Salipante, Classification and characterization of microsatellite instability across 18 cancer types, Nature medicine 22 (11) (2016) 1342–1350.

[2] C. R. Boland, A. Goel, Microsatellite instability in colorectal cancer, Gastroenterology 138 (6) (2010) 2073–2087.

[3] R. Murali, R. N. Grisham, R. A. Soslow, The roles of pathology in targeted therapy of women with gynecologic cancers, Gynecologic oncology 148 (1) (2018) 213–221.

[4] A. Cervantes, R. Adam, S. Roselló, D. Arnold, N. Normanno, J. Taïeb, J. Seligmann, T. De Baere, P. Osterlund, T. Yoshino, et al., Metastatic colorectal cancer: Esmo clinical practice guideline for diagnosis, treatment and follow-up, Annals of Oncology 34 (1) (2023) 10–32.

[5] D. T. Le, J. N. Durham, K. N. Smith, H. Wang, B. R. Bartlett, L. K. Aulakh, S. Lu, H. Kemberling, C. Wilt, B. S. Luber, et al., Mismatch repair deficiency predicts response of solid tumors to pd-1 blockade, Science 357 (6349) (2017) 409–413.

[6] A. Kavun, E. Veselovsky, A. Lebedeva, E. Belova, O. Kuznetsova, V. Yakushina, T. Grigoreva, V. Mileyko, M. Fedyanin, M. Ivanov, Microsatellite instability: a review of molecular epidemiology and implications for immune checkpoint inhibitor therapy, Cancers 15 (8) (2023) 2288.

[7] J. Ros, I. Baraibar, N. Saoudi, M. Rodriguez, F. Salvà, J. Tabernero, E. Élez, Immunotherapy for colorectal cancer with high microsatellite instability: the ongoing search for biomarkers, Cancers 15 (17) (2023) 4245.

[8] H. F. Berg, H. Engerud, M. Myrvold, H. E. Lien, M. E. Hjelmeland, M. K. Halle, K. Woie, E. A. Hoivik, I. S. Haldorsen, O. Vintermyr, et al., Mismatch repair markers in preoperative and operative endometrial cancer samples; expression concordance and prognostic value, British Journal of Cancer 128 (4) (2023) 647–655.

[9] E. Puliga, S. Corso, F. Pietrantonio, S. Giordano, Microsatellite instability in gastric cancer: Between lights and shadows, Cancer treatment reviews 95 (2021) 102175.

[10] C. Luchini, F. Bibeau, M. Ligtenberg, N. Singh, A. Nottegar, T. Bosse,R. Miller, N. Riaz, J.-Y. Douillard, F. Andre, et al., Esmo recommendations on microsatellite instability testing for immunotherapy in cancer, and its relationship with pd-1/pd-l1 expression and tumour mutational burden: a systematic review-based approach, Annals of Oncology 30 (8) (2019) 1232–1243.

[11] C.-W. Wang, H. Muzakky, N. P. Firdi, T.-C. Liu, P.-J. Lai, Y.-C. Wang, M.-H. Yu, T.-K. Chao, Deep learning to assess microsatellite instability directly from histopathological whole slide images in endometrial cancer, NPJ Digital Medicine 7 (1) (2024) 143.

[12] J. N. Kather, A. T. Pearson, N. Halama, D. Jäger, J. Krause, S. H. Loosen, A. Marx, P. Boor, F. Tacke, U. P. Neumann, et al., Deep learning can predict microsatellite instability directly from histology in gastrointestinal cancer, Nature medicine 25 (7) (2019) 1054–1056.

[13] J. N. Kather, L. R. Heij, H. I. Grabsch, C. Loeffler, A. Echle, H. S. Muti, J. Krause, J. M. Niehues, K. A. Sommer, P. Bankhead, et al., Pan-cancer image-based detection of clinically actionable genetic alterations, Nature cancer 1 (8) (2020) 789–799.

[14] M. Bilal, S. E. A. Raza, A. Azam, S. Graham, M. Ilyas, I. A. Cree, D. Snead, F. Minhas, N. M. Rajpoot, Development and validation of a weakly supervised deep learning framework to predict the status of molecular pathways and key mutations in colorectal cancer from routine histology images: a retrospective study, The Lancet Digital Health 3 (12) (2021) e763–e772.

[15] N. Zamanitajeddin, M. Jahanifar, M. Bilal, M. Eastwood, N. Rajpoot, Social network analysis of cell networks improves deep learning for prediction of molecular pathways and key mutations in colorectal cancer, Medical Image Analysis 93 (2024) 103071.

[16] H. Li, J. Qin, Z. Li, R. Ouyang, Z. Chen, S. Huang, S. Qin, Q. Huang, Systematic review and meta-analysis of deep learning for msi-h in colorectal cancer whole slide images, npj Digital Medicine 8 (1) (2025) 456.

[17] R. Yamashita, J. Long, T. Longacre, L. Peng, G. Berry, B. Martin, J. Higgins, D. L. Rubin, J. Shen, Deep learning model for the prediction of microsatellite instability in colorectal cancer: a diagnostic study, The Lancet Oncology 22 (2021) 132–141.

[18] A. Echle, H. I. Grabsch, P. Quirke, P. A. van den Brandt, N. P. West, G. G. Hutchins, L. R. Heij, X. Tan, S. D. Richman, J. Krause, et al., Clinical-grade detection of microsatellite instability in colorectal tumors by deep learning, Gastroenterology 159 (4) (2020) 1406–1416.

[19] X. Chang, J. Wang, G. Zhang, M. Yang, Y. Xi, C. Xi, G. Chen, X. Nie, B. Meng, X. Quan, Predicting colorectal cancer microsatellite instability with a self-attention-enabled convolutional neural network, Cell Reports Medicine 4 (2) (2023).

[20] S. Park, M. F. Pettigrew, Y. J. Cha, I.-H. Kim, M. Kim, I. Banerjee, Barnfather, J. R. Clemenceau, I. Jang, H. Kim, et al., Deep gaussian process with uncertainty estimation for microsatellite instability and immunotherapy response prediction from histology, npj Digital Medicine 8 (1) (2025) 294.

[21] M. Bilal, M. Raza, Y. Altherwy, A. Alsuhaibani, A. Abduljabbar, F. Almarshad, P. Golding, N. Rajpoot, et al., Foundation models in computational pathology: A review of challenges, opportunities, and impact, arXiv preprint arXiv:2502.08333 (2025).

[22] R. J. Chen, T. Ding, M. Y. Lu, D. F. Williamson, G. Jaume, A. H. Song, B. Chen, A. Zhang, D. Shao, M. Shaban, et al., Towards a general-purpose foundation model for computational pathology, Nature medicine 30 (3) (2024) 850–862.

[23] E. Vorontsov, A. Bozkurt, A. Casson, G. Shaikovski, M. Zelechowski, K. Severson, E. Zimmermann, J. Hall, N. Tenenholtz, N. Fusi, et al., A foundation model for clinical-grade computational pathology and rare cancers detection, Nature medicine 30 (10) (2024) 2924–2935.

[24] H. Xu, N. Usuyama, J. Bagga, S. Zhang, R. Rao, T. Naumann, C. Wong, Z. Gero, J. González, Y. Gu, et al., A whole-slide foundation model for digital pathology from real-world data, Nature 630 (8015) (2024) 181–188.

[25] M. Ilse, J. Tomczak, M. Welling, Attention-based deep multiple instance learning, in: International conference on machine learning, PMLR, 2018, pp. 2127–2136.

[26] Y. Wang, Y. G. Wang, C. Hu, M. Li, Y. Fan, N. Otter, I. Sam, H. Gou, Y. Hu, T. Kwok, et al., Cell graph neural networks enable the precise prediction of patient survival in gastric cancer, NPJ precision oncology 6 (1) (2022) 45.

[27] N. Zamanitajeddin, M. Jahanifar, N. Rajpoot, Cells are actors: Social network analysis with classical ml for sota histology image classification, in: International Conference on Medical Image Computing and Computer-Assisted Intervention, Springer, 2021, pp. 288–298.

[28] J. N. Weinstein, E. A. Collisson, G. B. Mills, K. R. Shaw, B. A. Ozenberger, K. Ellrott, I. Shmulevich, C. Sander, J. M. Stuart, The cancer genome atlas pan-cancer analysis project, Nature genetics 45 (10) (2013) 1113–1120.

[29] N. Zamanitajeddin, M. Jahanifar, K. Xu, F. Siraj, N. Rajpoot, Benchmarking domain generalization algorithms in computational pathology, arXiv preprint arXiv:2409.17063 (2024).

[30] M. Tan, Q. Le, Efficientnetv2: Smaller models and faster training, in: International conference on machine learning, PMLR, 2021, pp. 10096–10106.

[31] M. Jahanifar, M. Dawood, N. Zamanitajeddin, A. Shephard, B. S. Chohan, C. A. Bertram, N. Wahab, M. Eastwood, M. Aubreville, S. E. A. Raza, et al., Pan-cancer profiling of mitotic topology & mitotic errors: Insights into prognosis, genomic alterations, and immune landscape, medRxiv (2025) 2025–06.

[32] J. Pocock, S. Graham, Q. D. Vu, M. Jahanifar, S. Deshpande, G. Hadjigeorghiou, A. Shephard, R. M. S. Bashir, M. Bilal, W. Lu, et al., Tiatoolbox as an end-to-end library for advanced tissue image analytics, Communications medicine 2 (1) (2022) 1–14.

[33] Carmichael, A. H. Song, R. J. Chen, D. F. Williamson, T. Y. Chen, F. Mahmood, Incorporating intratumoral heterogeneity into weakly-supervised deep learning models via variance pooling, in: International Conference on Medical Image Computing and Computer-Assisted Intervention, Springer, 2022, pp. 387–397.

[34] S. Graham, Q. D. Vu, S. E. A. Raza, A. Azam, Y. W. Tsang, J. T. Kwak, N. Rajpoot, Hover-net: Simultaneous segmentation and classification of nuclei in multi-tissue histology images, Medical Image Analysis 58 (2019) 101563. doi:10.1016/j.media.2019.101563. URL https://www.sciencedirect.com/science/article/pii/S1361841519301045

[35] M. Jahanifar, A. Shephard, N. Zamanitajeddin, S. Graham, S. E. A. Raza, F. Minhas, N. Rajpoot, Mitosis detection, fast and slow: robust and efficient detection of mitotic figures, Medical Image Analysis 94 (2024) 103132.

[36] M. Newman, Networks, Oxford university press, 2018.

[37] H. B. Mann, D. R. Whitney, On a test of whether one of two random variables is stochastically larger than the other, The annals of mathematical statistics (1947) 50–60.

[38] Y. Benjamini, Y. Hochberg, Controlling the false discovery rate: a practical and powerful approach to multiple testing, Journal of the Royal statistical society: series B (Methodological) 57 (1) (1995) 289–300.

[39] N. Cliff, Dominance statistics: Ordinal analyses to answer ordinal questions., Psychological bulletin 114 (3) (1993) 494.

[40] T. Chen, C. Guestrin, Xgboost: A scalable tree boosting system, in: Proceedings of the 22nd acm sigkdd international conference on knowledge discovery and data mining, 2016, pp. 785–794.

[41] S. M. Lundberg, S.-I. Lee, A unified approach to interpreting model predictions, Advances in neural information processing systems 30 (2017).

[42] Z. Shao, H. Bian, Y. Chen, Y. Wang, J. Zhang, X. Ji, et al., Transmil: Transformer based correlated multiple instance learning for whole slide image classification, Advances in neural information processing systems 34 (2021) 2136–2147.

[43] B. Li, Y. Li, K. W. Eliceiri, Dual-stream multiple instance learning network for whole slide image classification with self-supervised contrastive learning, in: Proceedings of the IEEE/CVF conference on computer vision and pattern recognition, 2021, pp. 14318–14328.

[44] M. Y. Lu, D. F. Williamson, T. Y. Chen, R. J. Chen, M. Barbieri, F. Mahmood, Data-efficient and weakly supervised computational pathology on whole-slide images, Nature Biomedical Engineering 5 (6) (2021) 555–570.

[45] A. Su, H. Lee, M. Tran, R. C. dela Cruz, A. Sathe, X. Bai, I. Wichmann, L. Pflieger, B. Moulton, T. Barker, et al., The single-cell spatial landscape of stage iii colorectal cancers, NPJ Precision Oncology 9 (1) (2025) 101.

[46] G. Cerretelli, A. Ager, M. J. Arends, I. M. Frayling, Molecular pathology of lynch syndrome, The Journal of pathology 250 (5) (2020) 518–531.

[47] T. C. Smyrk, P. Watson, K. Kaul, H. T. Lynch, Tumor-infiltrating lymphocytes are a marker for microsatellite instability in colorectal carcinoma, Cancer 91 (12) (2001) 2417–2422.

[48] N. J. Llosa, M. Cruise, A. Tam, E. C. Wicks, E. M. Hechenbleikner, J. M. Taube, R. L. Blosser, H. Fan, H. Wang, B. S. Luber, et al., The vigorous immune microenvironment of microsatellite instable colon cancer is balanced by multiple counter-inhibitory checkpoints, Cancer discovery 5 (1) (2015) 43–51.

[49] S. Graham, F. Minhas, M. Bilal, M. Ali, Y. W. Tsang, M. Eastwood, N. Wahab, M. Jahanifar, E. Hero, K. Dodd, et al., Screening of normal endoscopic large bowel biopsies with interpretable graph learning: a retrospective study, Gut 72 (9) (2023) 1709–1721.

[50] A. Dosovitskiy, L. Beyer, A. Kolesnikov, D. Weissenborn, X. Zhai, T. Unterthiner, M. Dehghani, M. Minderer, G. Heigold, S. Gelly, et al., An image is worth 16×16 words: Transformers for image recognition at scale, arXiv preprint arXiv:2010.11929 (2020).

[51] A. Vaswani, N. Shazeer, N. Parmar, J. Uszkoreit, L. Jones, A. N. Gomez, L-. Kaiser, I. Polosukhin, Attention is all you need, Advances in neural information processing systems 30 (2017).

